# Sociodemographic disparities in knowledge, practices, and ability to comply with COVID-19 public health measures in Canada

**DOI:** 10.1101/2020.08.24.20180919

**Authors:** Gabrielle Brankston, Eric Merkley, David N. Fisman, Ashleigh R. Tuite, Zvonimir Poljak, Peter J. Loewen, Amy L. Greer

## Abstract

The effectiveness of public health interventions for mitigation of the coronavirus (COVID-19) pandemic depends on individual attitudes and the level of compliance toward these measures. We surveyed a representative sample of the Canadian population about risk perceptions, attitudes, and behaviours towards the Canadian COVID-19 public health response. Our analysis demonstrates that these risk perceptions, attitudes, and behaviours varied by several demographic variables identifying a number of areas in which policies could help address issues of public adherence. Examples include targeted messaging for men and younger age groups, social supports for those who need to self-isolate but may not have the means to do so, changes in workplace policies to discourage presenteeism, and provincially co-ordinated masking and safe school reopening policies. Taken together such measures are likely to mitigate the impact of the next pandemic wave in Canada.

## Introduction

The current coronavirus (COVID-19) pandemic represents a unique challenge for public health and health care systems. The virus is highly transmissible ^1,2^ and causes moderate to severe illness in approximately 20% of cases^3^. The first case of COVID-19 in Canada was reported on January 25, 2020 and was associated with international travel^4^. As of July 4, 2020, Canada has reported 106,842 cases and 8712 deaths^4^. The source of infection in 35% of these cases remains unknown^5^. In the absence of effective medical treatment options or a safe and effective vaccine, public health agencies have relied on non-pharmaceutical interventions (NPIs) to mitigate transmission of the virus. Physical distancing interventions act to reduce the person-to-person contact rate in a population thereby reducing the likelihood of disease transmission. All Canadian provinces and territories have instituted aggressive physical distancing measures in response to the COVID-19 pandemic including school closures, remote work, cancellation of mass gatherings, and the closure of all non-essential businesses. While these measures are expected to have slowed the rate of transmission of the virus in order to protect limited healthcare resources, they are disruptive to society, and the economy^6,7^.

Human behaviour is the main driver of respiratory disease transmission and in the absence of a vaccine or other pharmaceutical interventions, mitigation requires large scale behaviour change. As such, the effectiveness of public health interventions depends on the level of individual compliance. Perceived risk due to COVID-19 and attitudes toward these measures have a large impact on the willingness of people to make the behaviour changes necessary for public health measures to be effective^8^. As cases decline, provincial governments lift restrictions, and businesses and schools reopen, it is imperative that evidence is used to drive decision-making in order to minimize the transmission that is expected to occur. It is important to identify groups that are less likely to perceive COVID-19 as a risk, to perceive that public health measures are effective, and more likely to engage in behaviours associated with transmission of COVID-19. This information can be used to target messaging and develop policies to help support and encourage uptake of the necessary public health measures. The objectives of this study are to: 1) describe population attitudes and behaviours towards the Canadian COVID-19 public health response in May 2020, and 2) identify risk modifying behaviours based on demographic and household characteristics.

## Results

A total of 9,120 survey responses were received between May 7 and May 19, 2020. Survey responses were excluded from analysis if the survey was completed in less than 1/3 of the estimated completion time (n = 137), if the respondent reported their age as less than 18 years (n = 23), or if the survey was discontinued prior to completion for any reason including exceeding the age, gender, or province quotas (n = 3960). Respondents that completed the entire survey and were not screened out for any reason were included in the final sample resulting in 5000 high-quality survey responses.

A detailed description of the respondent population is included in the **Supplementary Materials (Table S1)**. For the 5000 surveys, the proportion of respondents living in each province, the male to female ratio, and the proportion of respondents in each age category were comparable to the 2016 Canadian Census of the population (data shown in **Table S1, Supplementary Materials**).

### Perceived Risk

**Table 1** describes respondents’ level of perceived risk as well as indicators of preparedness in the event of illness. Overall, 61.6% of respondents agreed that COVID-19 would be a serious illness for them, 21.6% agreed that they are likely to catch the virus, and 71.6% agreed that they are likely to transmit the virus if they did not follow public health advice. Perceived risk of serious illness due to COVID-19 increased with increasing age beyond 50 years however, perceived risk of contracting the virus was highest in the 30-39 year age group and decreased with increasing age up to the 60-69 year age group. Individuals who self-identified as being in a risk group were more likely to agree that they are likely to catch the virus and experience serious illness compared to other individuals, while those living with children under the age of 18 years or those in the paid workforce were less likely to agree that COVID-19 would be a serious illness for them compared with households containing no children or those not in the paid workforce, respectively. Risk perception was also associated with gender (**Table 1**).

**Table 1.** Indicators of perceived risk and preparedness in the event of illness stratified by sociodemographic characteristics. Values are reported as % (95% Confidence Interval), and those in bold font were statistically significant between subgroups (p < 0.0026). Cells denoted by “-“ signify that statistics were not run because the survey question was not relevant for one of the groups.

|  | Perceived Risk |  |  | Preparedness in the Event of Illness |  |  |  |
| --- | --- | --- | --- | --- | --- | --- | --- |
|  | Likely to contract COVID-19 (n = 5000) | Likely to be a serious illness for self (n = 5000) | Likely to transmit to others (n = 5000) | My co-workers would not expect me to work if ill (n = 2709) | I would still get paid (n = 2709) | I have enough food/supplies to last for 14 days (n = 5000) | Someone else would be able to look after my children (n = 938) |
| Overall | 21.6 %<br>(20.6 – 22.7) | 61.6%<br>(60.2 – 62.9) | 71.6 %<br>(70.3 – 72.8) | 50.9%<br>(49.0 – 52.8) | 51.1%<br>(49.2 – 53.0) | 70.8%<br>(69.6 – 72.1) | 59.8%<br>(56.7 – 63.0) |
| Gender |  |  |  |  |  |  |  |
| Women | 20.9%<br>(19.3 – 22.5) | 61.1%<br>(59.2 – 63.0) | **74.5%**<br>**(72.8 – 76.2)** | **55.8%**<br>**(53.1 – 58.5)** | 48.6%<br>(45.8 – 51.3) | **72.9%**<br>**(71.1 – 74.6)** | **53.2%**<br>**(48.8 – 57.7)** |
| Men | 22.3%<br>(20.6 – 23.9) | 62.0%<br>(60.1 – 63.9) | **68.5%**<br>**(66.6 – 70.3)** | **46.7%**<br>**(44.1 – 49.3)** | 53.5%<br>(50.9 – 56.1) | **68.9%**<br>**(67.0 – 70.7)** | **66.6%**<br>**(62.3 – 70.9)** |
| Age Group |  |  |  |  |  |  |  |
| 18-29 years | **25.8%**<br>**(22.8 – 28.9)** | **45.1%**<br>**(41.7 – 48.6)** | 71.0%<br>(67.8 – 74.2) | **40.1%**<br>**(35.7 – 44.5)** | **48.1%**<br>**(43.7 – 52.6)** | **62.5%**<br>**(59.1 – 65.9)** | **54.5%**<br>**(46.0 – 62.7)** |
| 30-39 years | **32.3%**<br>**(29.3 – 35.3)** | **51.8%**<br>**(48.6 – 55.0)** | 71.6%<br>(68.7 – 74.4) | **43.2%**<br>**(39.6 – 46.7)** | **55.4%**<br>**(51.9 – 58.9)** | **65.7%**<br>**(62.7 – 68.7)** | **57.1%**<br>**(52.0 – 62.2)** |
| 40-49 years | **24.3%**<br>**(21.3 – 27.3)** | **55.1%**<br>**(51.7 – 58.6)** | 70.3%<br>(67.1 – 73.4) | **52.5%**<br>**(48.6 – 56.4)** | **54.1%**<br>**(50.2 – 57.9)** | **66.3%**<br>**(63.0 – 69.6)** | **60.9%**<br>**(55.5 – 66.2)** |
| 50-59 years | **18.6%**<br>**(16.0 – 21.3)** | **62.7%**<br>**(59.5 – 66.0)** | 71.5%<br>(68.5 – 74.5) | **63.5%**<br>**(59.5 – 67.5)** | **51.0%**<br>**(46.8 – 55.2)** | **71.6%**<br>**(68.6 – 74.7)** | **78.0%**<br>**(69.5 – 86.5)** |
| 60-69 years | **13.4%**<br>**(11.3 – 15.6)** | **72.9%**<br>**(70.1 – 75.7)** | 70.6%<br>(67.8 – 73.5) | **62.0%**<br>**(55.9 – 68.2)** | **41.4%**<br>**(35.1 – 47.6)** | **77.8%**<br>**(75.2 – 80.4)** | **35.0%**<br>**(14.1 – 55.9)** |
| 70+ years | **13.8%**<br>**(11.1 – 16.4)** | **85.3%**<br>**(82.5 – 88.0)** | 75.6%<br>(72.3 – 78.9) | - | - | **82.5%**<br>**(79.5 – 85.4)** | - |
| Risk Group |  |  |  |  |  |  |  |
| Yes | **30.0%**<br>**(27.8 – 32.2)** | **86.1%**<br>**(84.4 – 87.8)** | 74.1%<br>(71.9 – 76.2) | **44.0%**<br>**(40.4 – 47.6)** | 52.9%<br>(49.3 – 56.5) | **73.8%**<br>**(71.7 – 75.9)** | 63.7%<br>(57.9 – 69.6) |
| No | **17.5%**<br>**(16.2 – 18.8)** | **49.6%**<br>**(48.0 – 51.3)** | 70.4%<br>(68.8 – 71.9) | **53.4%**<br>**(51.2 – 55.6)** | 50.5%<br>(48.3 – 52.7) | **69.4%**<br>**(67.8 – 70.9)** | 58.3%<br>(54.6 – 62.0) |
| Household children |  |  |  |  |  |  |  |
| Yes | **29.0%**<br>**(26.4 – 31.6)** | **56.0%**<br>**(53.1 – 58.9)** | 71.0%<br>(68.4 – 73.6) | 48.4%<br>(45.1 – 51.8) | 53.5%<br>(50.2 – 56.8) | **66.2%**<br>**(63.5 – 68.9)** | - |
| No | **19.4%**<br>**(18.2 – 20.7)** | **63.2%**<br>**(61.7 – 64.7)** | 71.8%<br>(70.3 – 73.2) | 52.0%<br>(49.7 – 54.3) | 50.0%<br>(47.7 – 52.3) | **72.2%**<br>**(70.8 – 73.6)** | - |
| Household Income |  |  |  |  |  |  |  |
| ≤ \$30,000 | 19.7%<br>(16.7 – 22.6) | 65.9%<br>(62.4 – 69.4) | 69.3%<br>(65.9 – 72.7) | 46.8%<br>(39.9 – 53.7) | **32.8%**<br>**(26.3 – 39.3)** | **61.8%**<br>**(58.2 – 65.4)** | 48.5%<br>(36.7 – 60.4) |
| > \$30,000 | 22.9%<br>(21.6 – 24.3) | 60.9%<br>(59.3 – 62.4) | 72.4%<br>(71.0 – 73.9) | 50.7%<br>(48.7 – 52.7) | **53.1%**<br>**(51.0 – 55.1)** | **72.4%**<br>**(71.0 – 73.9)** | 61.0%<br>(57.7 – 64.4) |
| Employment Status |  |  |  |  |  |  |  |
| Paid workforce | **26.3%**<br>**(24.7 – 28.0)** | **56.4%**<br>**(54.5 – 58.3)** | 70.7%<br>(68.9 – 72.4) | - | - | **68.6%**<br>**(67.0 – 70.5)** | 61.7%<br>(58.1 – 65.2) |
| Not in paid workforce | **15.7%**<br>**(14.2 – 17.3)** | **67.5%**<br>**(65.6 – 69.5)** | 72.6%<br>(70.7 – 74.4) | - | - | **73.5%**<br>**(71.6 – 75.3)** | 54.4%<br>(47.6 – 61.3) |
| Level of Education |  |  |  |  |  |  |  |
| Secondary or less | 18.5%<br>(16.6 – 21.0) | 61.8%<br>(59.1 – 64.6) | 71.1%<br>(68.6 – 73.7) | 53.6%<br>(48.9 – 58.2) | **40.3%**<br>**(35.7 – 44.9)** | 71.5%<br>(68.9 – 74.0) | 50.3%<br>(42.3 – 58.4) |
| College/trades or other qualification | 21.7%<br>(19.8 – 23.5) | 62.2%<br>(60.4 – 64.4) | 69.9%<br>(67.9 – 72.0) | 51.4%<br>(48.3 – 54.5) | **50.7%**<br>**(47.6 – 53.8)** | 71.3%<br>(69.3 – 73.3) | 58.5%<br>(53.4 – 63.7) |
| University degree | 23.3%<br>(21.4 – 25.2) | 60.7%<br>(58.5 – 62.9) | 73.5%<br>(71.5 – 75.5) | 49.5%<br>(46.8 – 52.3) | **55.2%**<br>**(52.5 – 57.9)** | 69.9%<br>(67.9 – 72.0) | 64.1%<br>(59.6 – 68.6) |

### Perceived Preparedness

A higher proportion of older individuals and women reported that co-workers would not expect them to work if sick (**Table 1**). Fewer individuals with no risk, and with higher incomes or education, thought that they would be expected to work while sick. Demographics also predicted confidence in access to a 14-day supply of food, and ability to find childcare. Males and more educated individuals had more confidence in ability to find childcare (**Table 1**).

### Perceived Effectiveness and Confidence in the Ability to Comply with Public Health Measures

At least 87% of respondents considered each of the public health measures described to be effective in reducing the transmission of COVID-19, with women and older individuals expressing greater faith in public health measures (**Table 2**). Those in the paid workforce were less likely to agree that each of the public health measures are effective except school closures, where there was no difference between groups.

**Table 2.**
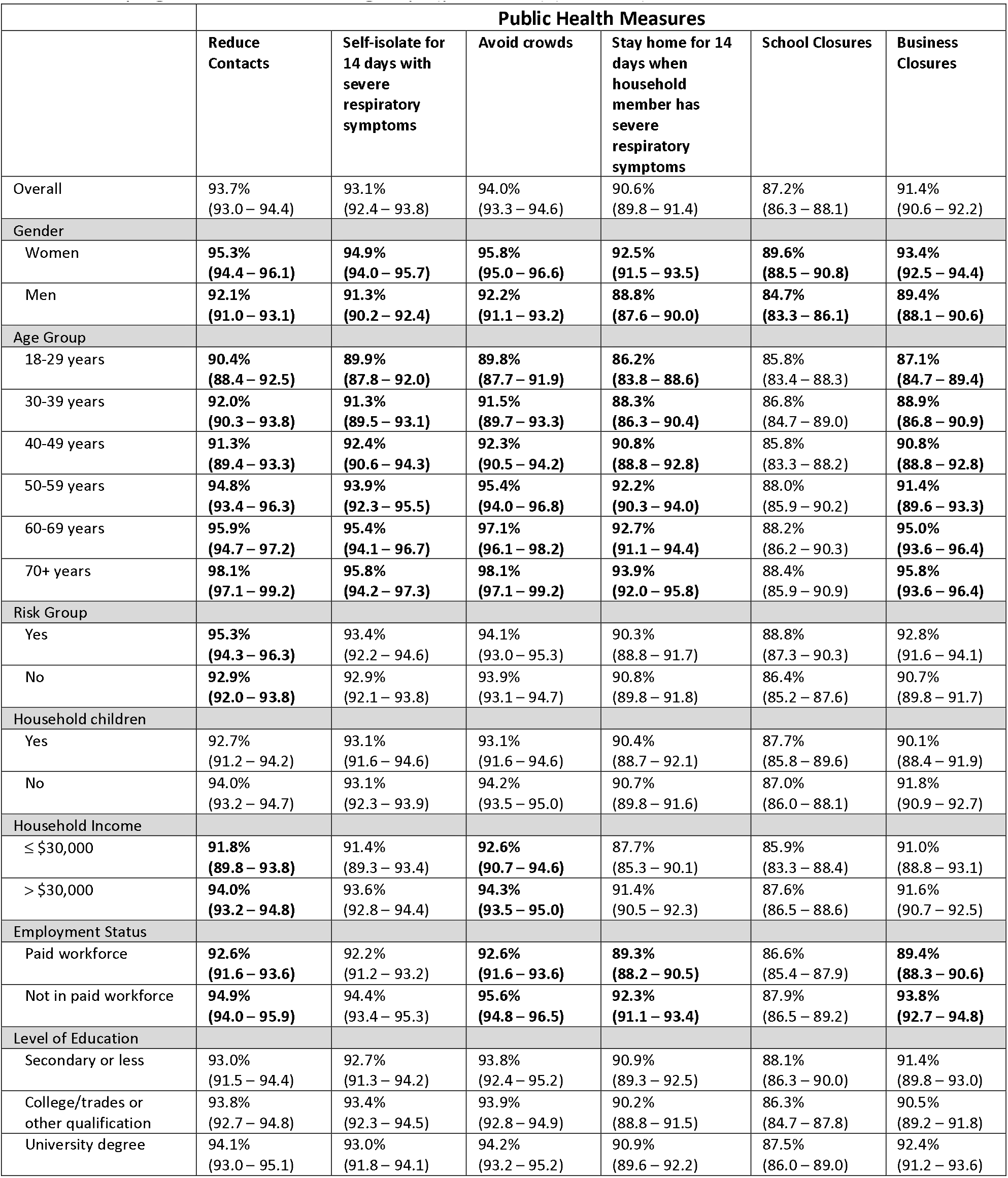
Perceived effectiveness of six different public health measures stratified by sociodemographic characteristics. Values are reported as % (95% Confidence Interval), and those in bold font were statistically significant between subgroups (p < 0.0026) (n = 5000).

|  | Public Health Measures |  |  |  |  |  |
| --- | --- | --- | --- | --- | --- | --- |
|  | Reduce Contacts | Self-isolate for 14 days with severe respiratory symptoms | Avoid crowds | Stay home for 14 days when household member has severe respiratory symptoms | School Closures | Business Closures |
| Overall | 93.7%<br>(93.0 – 94.4) | 93.1%<br>(92.4 – 93.8) | 94.0%<br>(93.3 – 94.6) | 90.6%<br>(89.8 – 91.4) | 87.2%<br>(86.3 – 88.1) | 91.4%<br>(90.6 – 92.2) |
| Gender |  |  |  |  |  |  |
| Women | **95.3%**<br>**(94.4 – 96.1)** | **94.9%**<br>**(94.0 – 95.7)** | **95.8%**<br>**(95.0 – 96.6)** | **92.5%**<br>**(91.5 – 93.5)** | **89.6%**<br>**(88.5 – 90.8)** | **93.4%**<br>**(92.5 – 94.4)** |
| Men | **92.1%**<br>**(91.0 – 93.1)** | **91.3%**<br>**(90.2 – 92.4)** | **92.2%**<br>**(91.1 – 93.2)** | **88.8%**<br>**(87.6 – 90.0)** | **84.7%**<br>**(83.3 – 86.1)** | **89.4%**<br>**(88.1 – 90.6)** |
| Age Group |  |  |  |  |  |  |
| 18-29 years | **90.4%**<br>**(88.4 – 92.5)** | **89.9%**<br>**(87.8 – 92.0)** | **89.8%**<br>**(87.7 – 91.9)** | **86.2%**<br>**(83.8 – 88.6)** | 85.8%<br>(83.4 – 88.3) | **87.1%**<br>**(84.7 – 89.4)** |
| 30-39 years | **92.0%**<br>**(90.3 – 93.8)** | **91.3%**<br>**(89.5 – 93.1)** | **91.5%**<br>**(89.7 – 93.3)** | **88.3%**<br>**(86.3 – 90.4)** | 86.8%<br>(84.7 – 89.0) | **88.9%**<br>**(86.8 – 90.9)** |
| 40-49 years | **91.3%**<br>**(89.4 – 93.3)** | **92.4%**<br>**(90.6 – 94.3)** | **92.3%**<br>**(90.5 – 94.2)** | **90.8%**<br>**(88.8 – 92.8)** | 85.8%<br>(83.3 – 88.2) | **90.8%**<br>**(88.8 – 92.8)** |
| 50-59 years | **94.8%**<br>**(93.4 – 96.3)** | **93.9%**<br>**(92.3 – 95.5)** | **95.4%**<br>**(94.0 – 96.8)** | **92.2%**<br>**(90.3 – 94.0)** | 88.0%<br>(85.9 – 90.2) | **91.4%**<br>**(89.6 – 93.3)** |
| 60-69 years | **95.9%**<br>**(94.7 – 97.2)** | **95.4%**<br>**(94.1 – 96.7)** | **97.1%**<br>**(96.1 – 98.2)** | **92.7%**<br>**(91.1 – 94.4)** | 88.2%<br>(86.2 – 90.3) | **95.0%**<br>**(93.6 – 96.4)** |
| 70+ years | **98.1%**<br>**(97.1 – 99.2)** | **95.8%**<br>**(94.2 – 97.3)** | **98.1%**<br>**(97.1 – 99.2)** | **93.9%**<br>**(92.0 – 95.8)** | 88.4%<br>(85.9 – 90.9) | **95.8%**<br>**(93.6 – 96.4)** |
| Risk Group |  |  |  |  |  |  |
| Yes | **95.3%**<br>**(94.3 – 96.3)** | 93.4%<br>(92.2 – 94.6) | 94.1%<br>(93.0 – 95.3) | 90.3%<br>(88.8 – 91.7) | 88.8%<br>(87.3 – 90.3) | 92.8%<br>(91.6 – 94.1) |
| No | **92.9%**<br>**(92.0 – 93.8)** | 92.9%<br>(92.1 – 93.8) | 93.9%<br>(93.1 – 94.7) | 90.8%<br>(89.8 – 91.8) | 86.4%<br>(85.2 – 87.6) | 90.7%<br>(89.8 – 91.7) |
| Household children |  |  |  |  |  |  |
| Yes | 92.7%<br>(91.2 – 94.2) | 93.1%<br>(91.6 – 94.6) | 93.1%<br>(91.6 – 94.6) | 90.4%<br>(88.7 – 92.1) | 87.7%<br>(85.8 – 89.6) | 90.1%<br>(88.4 – 91.9) |
| No | 94.0%<br>(93.2 – 94.7) | 93.1%<br>(92.3 – 93.9) | 94.2%<br>(93.5 – 95.0) | 90.7%<br>(89.8 – 91.6) | 87.0%<br>(86.0 – 88.1) | 91.8%<br>(90.9 – 92.7) |
| Household Income |  |  |  |  |  |  |
| ≤ \$30,000 | **91.8%**<br>**(89.8 – 93.8)** | 91.4%<br>(89.3 – 93.4) | **92.6%**<br>**(90.7 – 94.6)** | 87.7%<br>(85.3 – 90.1) | 85.9%<br>(83.3 – 88.4) | 91.0%<br>(88.8 – 93.1) |
| > \$30,000 | **94.0%**<br>**(93.2 – 94.8)** | 93.6%<br>(92.8 – 94.4) | **94.3%**<br>**(93.5 – 95.0)** | 91.4%<br>(90.5 – 92.3) | 87.6%<br>(86.5 – 88.6) | 91.6%<br>(90.7 – 92.5) |
| Employment Status |  |  |  |  |  |  |
| Paid workforce | **92.6%**<br>**(91.6 – 93.6)** | 92.2%<br>(91.2 – 93.2) | **92.6%**<br>**(91.6 – 93.6)** | **89.3%**<br>**(88.2 – 90.5)** | 86.6%<br>(85.4 – 87.9) | **89.4%**<br>**(88.3 – 90.6)** |
| Not in paid workforce | **94.9%**<br>**(94.0 – 95.9)** | 94.4%<br>(93.4 – 95.3) | **95.6%**<br>**(94.8 – 96.5)** | **92.3%**<br>**(91.1 – 93.4)** | 87.9%<br>(86.5 – 89.2) | **93.8%**<br>**(92.7 – 94.8)** |
| Level of Education |  |  |  |  |  |  |
| Secondary or less | 93.0%<br>(91.5 – 94.4) | 92.7%<br>(91.3 – 94.2) | 93.8%<br>(92.4 – 95.2) | 90.9%<br>(89.3 – 92.5) | 88.1%<br>(86.3 – 90.0) | 91.4%<br>(89.8 – 93.0) |
| College/trades or other qualification | 93.8%<br>(92.7 – 94.8) | 93.4%<br>(92.3 – 94.5) | 93.9%<br>(92.8 – 94.9) | 90.2%<br>(88.8 – 91.5) | 86.3%<br>(84.7 – 87.8) | 90.5%<br>(89.2 – 91.8) |
| University degree | 94.1%<br>(93.0 – 95.1) | 93.0%<br>(91.8 – 94.1) | 94.2%<br>(93.2 – 95.2) | 90.9%<br>(89.6 – 92.2) | 87.5%<br>(86.0 – 89.0) | 92.4%<br>(91.2 – 93.6) |

More than 90% of respondents reported that they were confident in their ability to comply with each of the five public health measures (**Table 3**), with greater confidence on most measures in women and older individuals. Lower-income individuals were less confident in their ability to avoid public transportation and comply with quarantine. Less confidence was seen in the paid workforce, compared to those who were unemployed, retired, or working within the home.

**Table 3.** Confidence in the ability to comply with five different public health measures stratified by sociodemographic characteristics. Values are reported as % (95% Confidence Interval), and those in bold font were statistically significant between subgroups (p < 0.0026) (n = 5000).

|  | Public Health Measures |  |  |  |  |
| --- | --- | --- | --- | --- | --- |
|  | Reduce Contacts | Self-isolate for 14 days with severe respiratory symptoms | Avoid crowds | Stay home for 14 days when household member has severe respiratory symptoms | Avoid public transportation |
| Overall | 93.7%<br>(93.0 – 94.4) | 93.0%<br>(92.3 – 93.7) | 93.3%<br>(92.6 – 94.0) | 91.0%<br>(90.2 – 91.8) | 90.0%<br>(90.1 – 91.7) |
| **Gender** |  |  |  |  |  |
| Women | 94.6%<br>(93.7 – 95.5) | **94.9%**<br>**(94.1 – 95.8)** | **94.7%**<br>**(93.8 – 95.5)** | **92.7%**<br>**(91.6 – 93.7)** | **92.9%**<br>**(91.9 – 93.9)** |
| Men | 92.9%<br>(91.9 – 93.9) | **91.1%**<br>**(90.0 – 92.2)** | **91.9%**<br>**(90.8 – 92.9)** | **89.3%**<br>**(88.1 – 90.6)** | **88.9%**<br>**(87.6 – 90.1)** |
| **Age Group** |  |  |  |  |  |
| 18-29 years | **90.2%**<br>**(88.1 – 92.2)** | **87.7%**<br>**(85.4 – 90.0)** | **88.1%**<br>**(85.8 – 90.4)** | **85.6%**<br>**(83.2 – 88.1)** | **85.3%**<br>**(82.8 – 87.8)** |
| 30-39 years | **92.7%**<br>**(91.0 – 94.3)** | **90.7%**<br>**(88.8 – 92.5)** | **89.2%**<br>**(87.2 – 91.2)** | **87.6%**<br>**(85.5 – 89.7)** | **86.9%**<br>**(84.8 – 89.1)** |
| 40-49 years | **92.1%**<br>**(90.2 – 93.9)** | **92.2%**<br>**(90.3 – 94.0)** | **91.7%**<br>**(89.8 – 93.6)** | **89.5%**<br>**(87.5 – 91.7)** | **89.3%**<br>**(87.2 – 91.5)** |
| 50-59 years | **94.4%**<br>**(92.8 – 95.9)** | **94.3%**<br>**(92.7 – 95.8)** | **95.7%**<br>**(94.3 – 97.0)** | **93.9%**<br>**(92.3 – 95.5)** | **92.2%**<br>**(90.3 – 94.0)** |
| 60-69 years | **96.7%**<br>**(95.6 – 97.8)** | **96.4%**<br>**(95.3 – 97.6)** | **97.2%**<br>**(96.2 – 98.3)** | **94.2%**<br>**(92.7 – 95.6)** | **95.0%**<br>**(93.6 – 95.4)** |
| 70+ years | **96.1%**<br>**(94.6 – 97.6)** | **97.2%**<br>**(95.9 – 98.5)** | **98.4%**<br>**(97.5 – 99.4)** | **95.3%**<br>**(93.7 – 97.0)** | **97.3%**<br>**(96.1 – 98.6)** |
| **Risk Group** |  |  |  |  |  |
| Yes | 95.1%<br>(94.1 – 96.2) | 92.8%<br>(91.6 – 94.1) | 94.2%<br>(93.1 – 95.3) | 91.0%<br>(89.6 – 92.4) | 91.5%<br>(90.1 – 92.9) |
| No | 93.0%<br>(92.2 – 93.9) | 93.1%<br>(92.3 – 94.0) | 92.8%<br>(92.0 – 93.7) | 90.9%<br>(90.0 – 91.9) | 90.6%<br>(89.6 – 91.5) |
| **Household children** |  |  |  |  |  |
| Yes | 92.7%<br>(91.2 – 94.2) | 91.7%<br>(90.1 – 93.3) | 91.7%<br>(90.1 – 93.3) | 90.0%<br>(88.3 – 91.8) | 89.2%<br>(87.4 – 91.0) |
| No | 94.0%<br>(93.3 – 94.8) | 93.4%<br>(92.6 – 94.2) | 93.8%<br>(93.0 – 94.5) | 91.2%<br>(90.3 – 92.1) | 91.4%<br>(90.5 – 92.3) |
| **Household Income** |  |  |  |  |  |
| ≤ \$30,000 | 92.2%<br>(90.3 – 94.2) | 91.0%<br>(88.8 – 93.1) | 92.2%<br>(90.3 – 94.2) | **88.1%**<br>**(85.7 – 90.5)** | **86.4%**<br>**(83.9 – 89.0)** |
| > \$30,000 | 94.2%<br>(93.4 – 94.9) | 93.5%<br>(92.7 – 94.3) | 93.6%<br>(92.8 – 94.4) | **91.4%**<br>**(90.5 – 92.3)** | **91.8%**<br>**(90.9 – 92.7)** |
| **Employment Status** |  |  |  |  |  |
| Paid workforce | **92.4%**<br>**(91.4 – 93.4)** | **91.4%**<br>**(90.3 – 92.5)** | **91.1%**<br>**(90.1 – 92.2)** | **88.9%**<br>**(87.7 – 90.1)** | **88.9%**<br>**(87.7 – 90.1)** |
| Not in paid workforce | **95.3%**<br>**(94.4 – 96.2)** | **94.9%**<br>**(94.0 – 95.9)** | **95.9%**<br>**(95.1 – 96.7)** | **93.4%**<br>**(92.4 – 94.5)** | **93.1%**<br>**(92.0 – 94.1)** |
| **Level of Education** |  |  |  |  |  |
| Secondary or less | 92.7%<br>(91.3 – 94.2) | 92.5%<br>(91.0 – 94.0) | 93.1%<br>(91.7 – 94.6) | 90.1%<br>(88.4 – 91.8) | 90.4%<br>(88.7 – 92.1) |
| College/trades or other qualification | 93.6%<br>(92.5 – 94.7) | 93.6%<br>(92.5 – 94.7) | 93.7%<br>(92.6 – 94.8) | 92.2%<br>(91.0 – 93.4) | 91.4%<br>(90.2 – 92.7) |
| University degree | 94.4%<br>(93.4 – 95.5) | 92.8%<br>(91.6 – 94.0) | 93.0%<br>(91.8 – 94.1) | 90.3%<br>(88.9 – 91.6) | 90.6%<br>(89.3 – 91.9) |

### Childcare

Respondents with household members who were 14 years of age or younger were asked about childcare provision when schools and daycares were closed due to the pandemic (n = 938). More than 80% of respondents reported that a parent provided childcare for their children during this time **(Figure 1A)**. Only 12.1% (95% CI: 10.0% - 14.1%) of those requiring childcare used providers that were not part of their household. Of the parents providing childcare, parents in the workforce provided the greatest proportion of childcare duties (52%) **(Figure IB)**.

**Figure 1.**
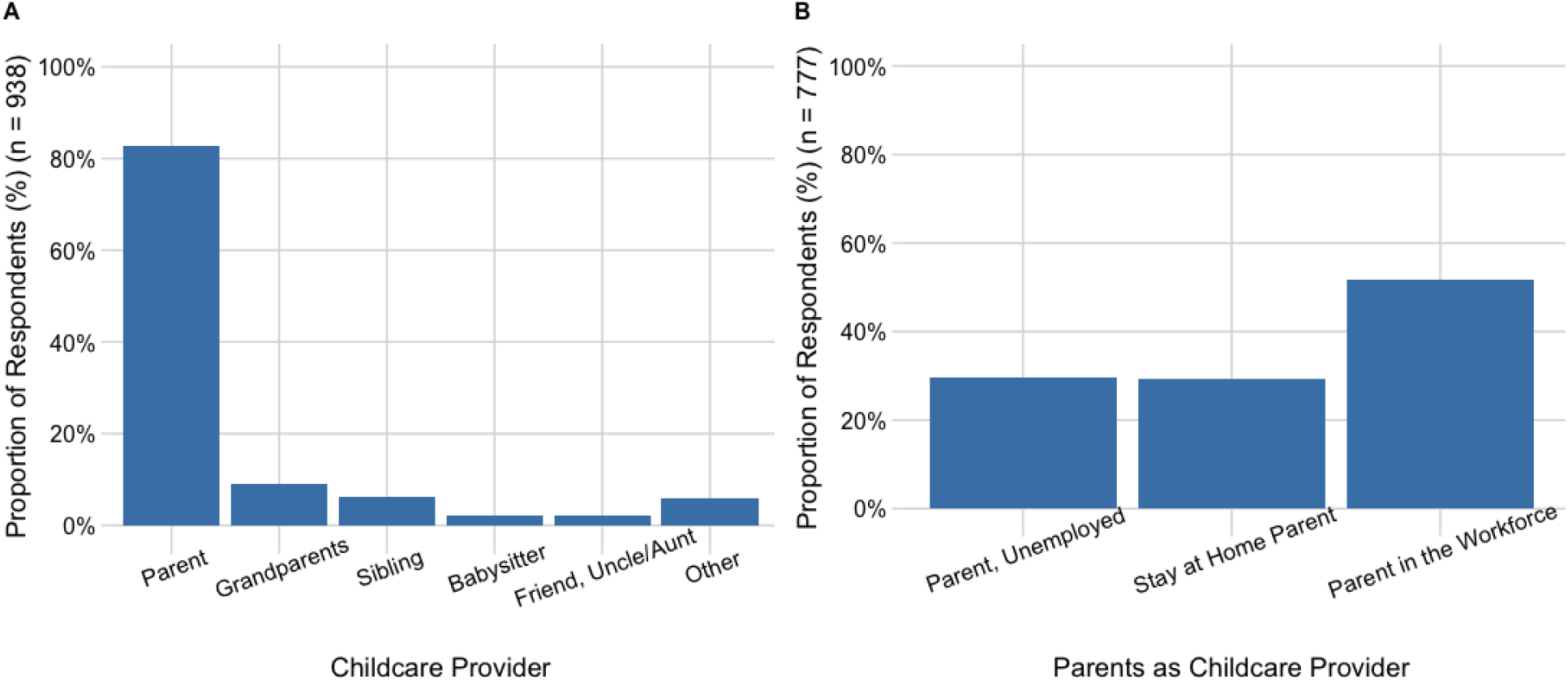
**Panel A**. Respondents with children under the age of 14 years (n = 938) reported on which individuals looked after the children in their household during school and daycare closures due to the pandemic. **Panel B**. Respondents who reported that parents provided childcare during school and daycare closures (n = 777), also identified the employment circumstances of the parent who provided the childcare. The category ‘Parent in the Workforce’ includes those who were working remotely, working part-time, those who took leave from their job, or those who were unemployed due to COVID-19 but otherwise have been working.

### Predictors of Mask Use

The proportion of respondents who wore a mask in the 24 hours prior to survey completion was 32.4% (95% CI: 31.1% – 33.7%) for an average duration of 96.5 (SD 412) minutes. Respondents from Ontario (where physical distancing measures were still in place at the time of the survey) reported the highest level of mask use while those from Prince Edward Island (where physical distancing recommendations were beginning to relax at the time of the survey) reported the lowest mask use (**Figure 2A**). The most common locations to wear a mask were in supermarkets or other stores, anywhere outside the home, and walking on the street (**Figure 2B**); 41.7% (95% CI: 34.4% - 49.0%) of mask-wearing transit-users had worn a mask on transit in the past 24 hours.

**Figure 2.**
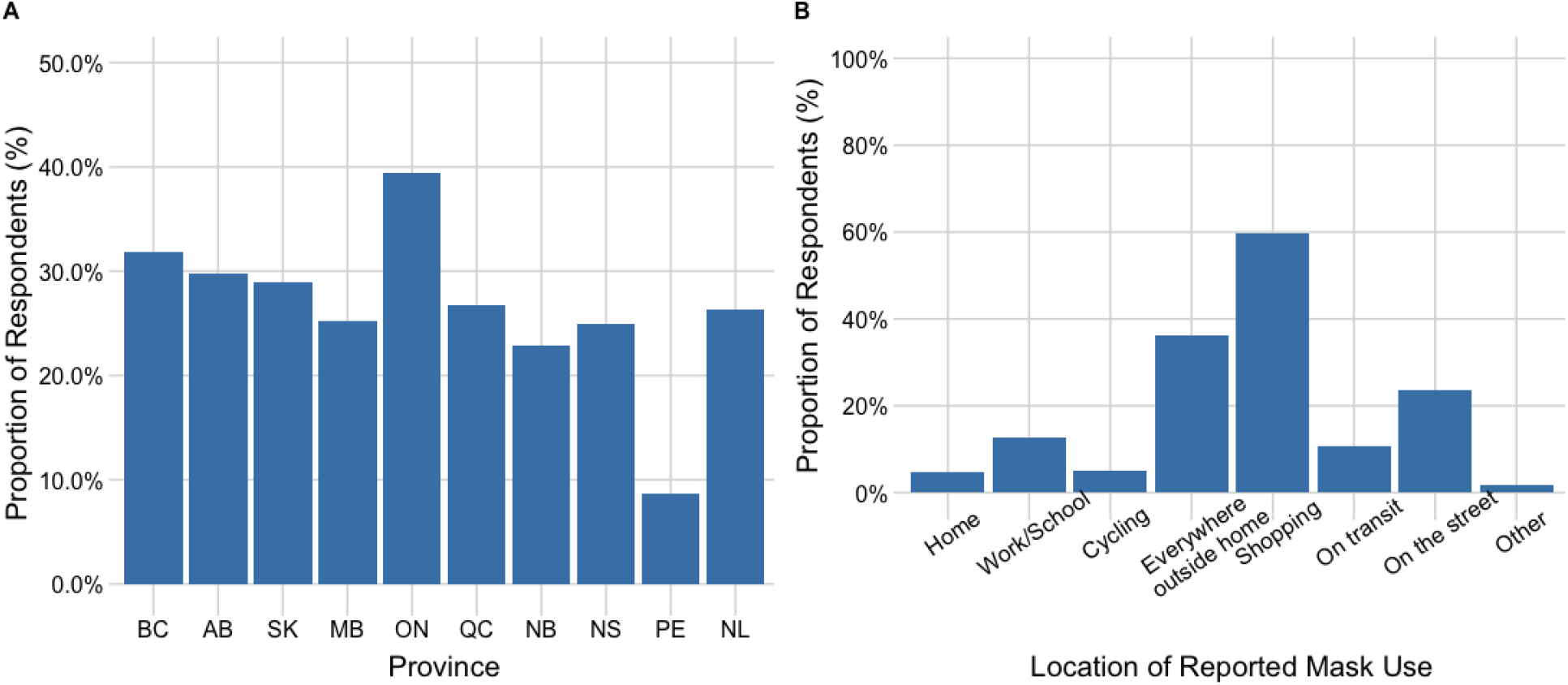
Respondents were asked if they had worn a face mask in the 24 hours prior to survey completion. **Panel A** represents reported mask use by province of residence. **Panel B** identifies the location(s) of mask use for respondents who reported wearing a mask in the previous 24 hours (n = 1622).

Factors associated with mask use are shown in **Table 4**; mask use was increased in households with more than one adult, with children, or with multiple generations; and in individuals with university education, or reporting that they would be at risk of serious illness with COVID-19 or at increased risk of developing COVID-19. An interaction was found between age and high-risk conditions with younger (< 40 years), high risk individuals more likely to have reported mask use compared with 40-49-year-old respondents who were not in a risk group.

**Table 4.** Results of a logistic regression assessing factors associated with mask use in the 24 hours prior to survey completion. Values are reported as adjusted odds ratios (95% confidence interval) and those in bold font were statistically significant (p < 0.05) (n = 5000).

| Variable | Adjusted OR (95% CI) | P Wald's test | P (L-R test) |
| --- | --- | --- | --- |
| **Household composition** |  |  | 0.004 |
| Single person living alone (referent) |  |  |  |
| **Adults only living together** | **1.33 (1.13 - 1.55)** | **< 0.001** |  |
| **Family with children** | **1.25 (1.02 - 1.53)** | **0.03** |  |
| **>2 generations living together** | **1.72 (1.12 - 2.65)** | **0.01** |  |
| Grandparents living with their grandchildren only | 1.16 (0.28 - 4.73) | 0.83 |  |
| **Age Category** |  |  | 1 |
| **18-29 years** | **1.62 (1.24 - 2.11)** | **< 0.001** |  |
| 30-39 years | 1.22 (0.95 - 1.56) | 0.12 |  |
| 40-49 years (referent) | - | - |  |
| 50-59 years | 1.23 (0.94 - 1.61) | 0.13 |  |
| **60-69 years** | **1.45 (1.09 - 1.94)** | **0.01** |  |
| Over 70 years | 1.41 (0.99 - 2.01) | 0.06 |  |
| **Respondent risk group** |  |  | 1 |
| **Respondent is in a high risk group** | **1.48 (1.04 - 2.11)** | **0.03** |  |
| **Size of the geographic region of residence** |  |  | < 0.001 |
| Large city (referent) |  |  |  |
| **Medium sized city** | **0.73 (0.63 - 0.85)** | **< 0.001** |  |
| **Large town** | **0.60 (0.48 - 0.76)** | **< 0.001** |  |
| **Small town** | **0.53 (0.43 - 0.65)** | **< 0.001** |  |
| **Rural place** | **0.38 (0.29 - 0.50)** | **< 0.001** |  |
| **Education level of respondent** |  |  | 0.02 |
| Secondary or less (referent) |  |  |  |
| College/Trade/Other qualification | 1.00 (0.85 - 1.18) | 1.0 |  |
| **University (Bachelor degree or higher)** | **1.2 (1.02 - 1.43)** | **0.03** |  |
| **Employment status of respondent** |  |  | < 0.001 |
| Unemployed, Student, Retired, Work w/in Home (referent) |  |  |  |
| **Employed FT, PT, Self Employed** | **1.33 (1.14 - 1.55)** | **< 0.001** |  |
| **Perceived risk of contracting the virus** |  |  | < 0.001 |
| **Likely to contact the virus** | **1.31 (1.12 - 1.52)** | **< 0.001** |  |
| **Perceived risk of serious illness due to COVID-19** |  |  | < 0.001 |
| **COVID-19 would be a serious illness for respondent** | **1.61 (1.39 - 1.85)** | **< 0.001** |  |
| **Interaction between age category and respondent risk group** |  |  | < 0.001 |
| **18-29 years in a risk group** | **1.75 (1.06 - 2.90)** | **0.03** |  |
| **30-39 years in a risk group** | **1.62 (1.01 - 2.59)** | **0.05** |  |
| 40-49 years, not high risk (referent) | - | - |  |
| 50-59 years in a risk group | 0.87 (0.54 - 1.39) | 0.56 |  |
| 60-69 years in a risk group | 0.73 (0.47 - 1.15) | 0.18 |  |
| Over 70 years in a risk group | 0.90 (0.55 - 1.48) | 0.68 |  |

### Direct contact with non-household members

The proportion of respondents who had engaged in an activity with non-household contacts in the 7 days prior to survey completion was 24.4% (95% CI: 23.2% - 25.6%) (**Figure 3A**). More non-household contacts were reported for provinces which were more advanced in the de-escalation of physical distancing (e.g. PEI) at the time of survey completion however, in provinces where physical distancing was still in place during the survey period (e.g. ON), approximately 20% of respondents were reporting non-household contacts in May 2020. Of the respondents who reported non-household contacts, 62% reported that this occurred once or twice in a seven-day period while almost 23% reported having non-household contacts more than 3 days out of the 7-day period prior to survey completion (**Figure 3B**). Younger individuals and individuals with higher income were more likely to have participated in an activity with someone outside their household (**Table 5**) compared with older respondents and those who reported no income at all, respectively. Respondents who reported a household income of more than $110,000 were 2.65 (95% CI: 1.25 – 5.62) to 3.57 (95% CI: 1.59 – 6.07) times as likely to have participated in an activity with someone outside of their household. Perceived risk of serious illness and belief in the effectiveness of reducing contact numbers were associated with less interaction with individuals outside the household.

**Figure 3.**
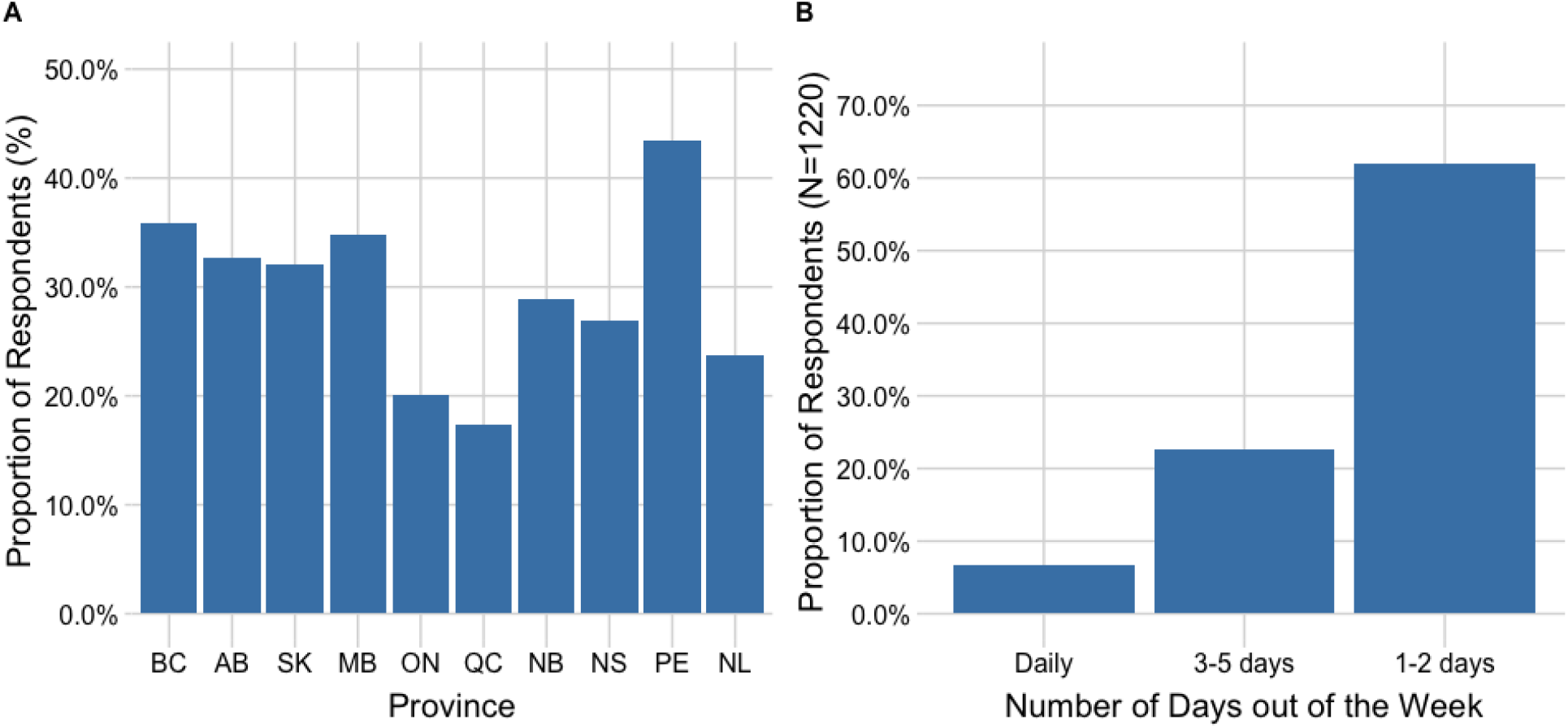
**Panel A**. Proportion of respondents reporting contact with non-household members in the 7 days prior to survey completion. More non-household contacts were reported for provinces which were more advanced in the de-escalation of physical distancing (e.g. PEI) at the time of survey completion however, in provinces where physical distancing was still in place during the survey period (e.g. ON), approximately 20% of respondents were reporting non-household contacts in May 2020. **Panel B**. The number of days in the past week respondents engaged in an activity with a non-household contact, for those reporting such activity (n = 1220).

**Table 5.**
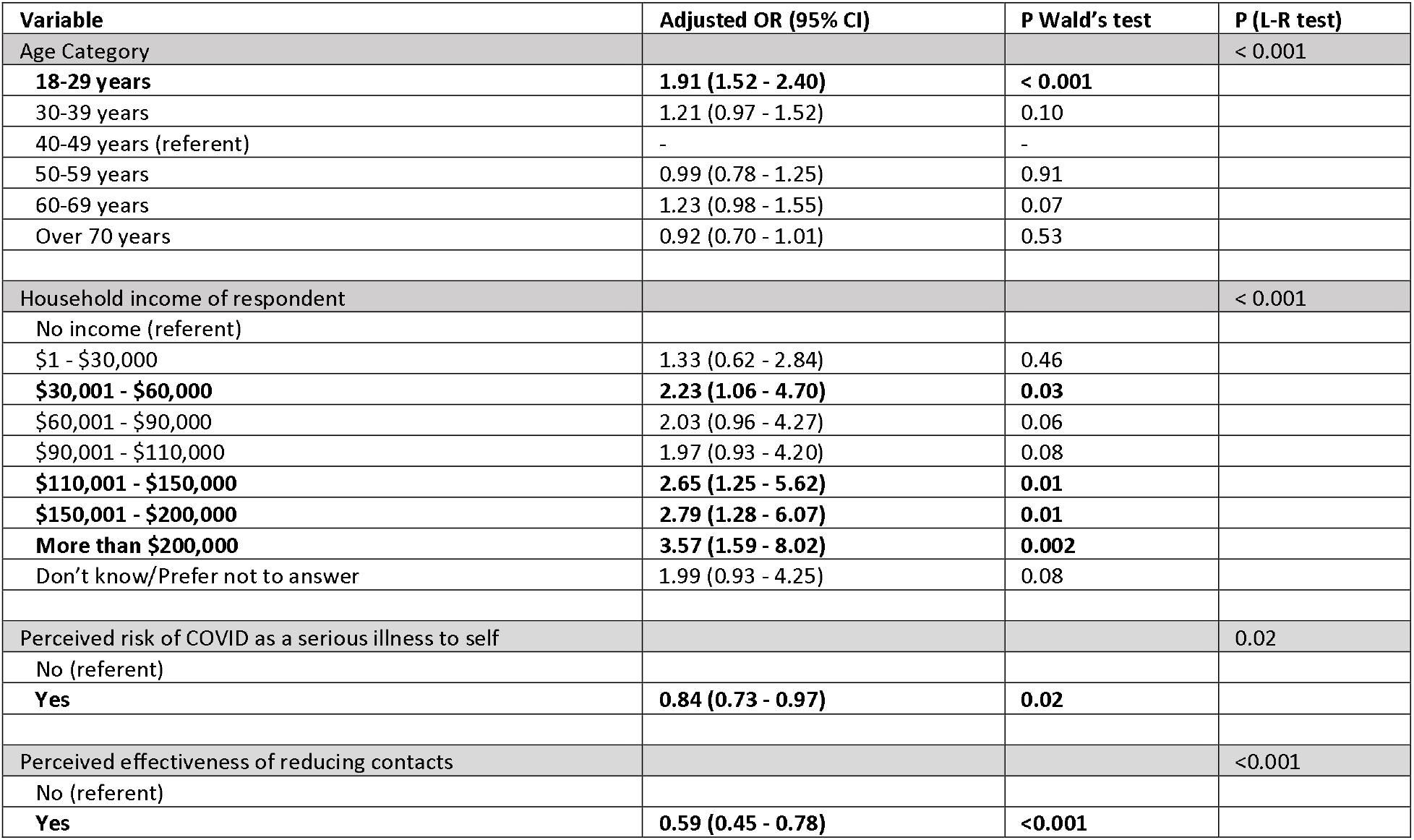
Results of a logistic regression assessing factors associated with engaging in an activity with non-household contacts in the 7 days prior to survey completion. Values are reported as adjusted odds ratios (95% Confidence Interval) and those in bold font are statistically significant (p < 0.05) (n = 4784).

## Discussion

At the time of data collection, Canadian provinces were in various stages of reopening the economy. If our collective priority is to reopen the economy and maintain that status, we need to ensure that individuals comply with public health measures that prevent and control the transmission of COVID-19. The results from this study have identified a number of areas in which policies could help address issues of public adherence.

Individuals need to feel supported in complying with public health measures. Our findings of reduced confidence in ability to comply with public health measures are consistent with other research demonstrating that those with a low income^9^ and those in younger age groups ^10^ are less prepared in the event of illness. Compliance in the event of self-isolation or quarantine is at least partially dependent on preparedness; there is a need to develop supports for those who need to self-isolate but may not have the means to do so. We also found evidence that individuals with fewer resources would be at risk of presenteeism (attending work while sick) due to co-worker expectations. Presenteeism risk was gendered, with fewer women than men anticipating that co-workers would expect them work even when sick. More individuals with less education or income were at risk of not being paid if they took sick leave. Presenteeism has been shown to be prevalent among occupations with high contact rates, including the care, welfare, and education sectors ^11,12^. Determinants of presenteeism include job insecurity, workplace performance indicators that include attendance rates, and limited entitlement to paid sick leave ^13^. Investigators in Israel demonstrated that paid sick time increases compliance with stay-home-when-sick policies from 57% with no compensation to almost 100% when compensation was assumed ^14^. These findings highlight the need for a shift in workplace culture toward discouraging presenteeism and ensuring paid sick time.

Our finding that the majority of respondents with dependent children were responsible for childcare at the same time as maintaining employment when schools and daycares were closed due to the pandemic highlights the need for provincially co-ordinated plans for safe school reopening. Statistics Canada estimates that there are more than 10 million families with children living in Canada ^15^ and almost 70% of families with dependent children have two employed parents ^16^. School boards have a variety of scenarios for reopening schools, some of which include staggering the days in which children will attend in person. These return-to-school scenarios are not well-defined in many jurisdictions, creating a childcare dilemma for parents who need to work.

We found that both perceived ability to comply with public health measures, and perceived effectiveness of such measures, varied by age and gender, emphasizing the need for targeted messaging ^8,17–24^. The finding that perceived risk of serious illness increased with age group is consistent with past research^25–28^ and is in line with empiric estimates of illness risk in older individuals^29^. Perceived lack of risk in younger individuals was associated with poor compliance with public health measures and is consistent with a growing body of evidence demonstrating that male gender and younger age groups engage in more COVID-19 risk behaviours^22,24,30–32^. Younger adults tend to have larger contact networks than older adults ^33^ which likely partially explains these results. Recent increases in cases of COVID-19 in adolescents and young adults have been attributed to greater mixing amongst this age group combined with lower adherence to physical distancing measures^34^.

The evidence for the efficacy of non-medical masks for COVID-19 prevention continues to grow^35–38^. While the survey question for mask use was not restricted to people who had left their household in the previous 24 hours, fewer than one-third of respondents reported wearing a face mask in the 24 hours prior to survey completion. Mask use was associated with household composition and the strongest association was belonging to a household with more than two generations living together, likely reflecting concern for the safety of older individuals in households. Increased mask use in the youngest age group may reflect younger individuals working in essential service jobs at the time of the survey (e.g. grocery stores). As with other preventive measures, compliance with masks was more likely in individuals with greater self-perceived risk.

### Limitations

While every effort was made to ensure representativeness of the study population, we note several potential biases, including non-representativeness of the sample (a risk with any survey), the online nature of the survey, which limits participation to those who use the Internet, and self-report which introduces the potential for recall, response, and social desirability biases. The large sample size means that statistical significance is seen with small absolute differences. Finally, knowledge about COVID-19 and recommended behaviours is changing rapidly. These data were collected in May 2020 during a time in which provinces were in different phases of public health de-escalation and indoor masking orders were not widespread, so these data are best interpreted as a snapshot in time.

## Conclusion

The results of this study highlight the need for the development of enhanced messaging in order to further support improved compliance with public health measures, including masking. Work is needed to identify strategies and develop tools for targeted messaging to groups that are more likely to engage in risk behaviours, and social support is needed for lower income individuals to enable periods of self-isolation and childcare should they become ill, to permit school opening, and to discourage presenteeism. Taken together such measures are likely to mitigate the impact of the next pandemic wave in Canada.

## Methods

### Data collection

The study protocol was approved by the University of Guelph Research Ethics Board (protocol #20-04-011) and the University of Toronto Research Ethics Board (protocol #38251). The research company, Dynata, was contracted to conduct an electronic survey of Canadian adults between May 7 and May 19, 2020. Dynata recruited individuals from their panel of survey respondents. Participants were paid a nominal amount for completing the survey. Informed consent was obtained prior to survey completion by providing information about the study, ensuring anonymity and confidentiality, and providing the process to withdraw from the survey. Respondents provided informed consent after reading the study information by choosing to continue to the survey questions. Representativeness of the survey sample population was ensured by setting quotas on age, gender, language, and region of residence (i.e., Atlantic, Quebec, Ontario, and West) based on 2016 Canadian Census data ^39^. Enrollment into the survey within each stratum was on a first-come, first-served basis.

The survey instrument was adapted from Jarvis et al^40^ and posed questions about self-perceived risk of COVID-19 infection, as well as attitudes and behaviours regarding COVID-19 public health measures. These included questions related to adherence to physical distancing recommendations. Participants provided information about their age, gender, province of residence, education level, employment status, household composition, household income, and the general size of their location of residence with options ranging from large city to rural. Participants were also asked whether they would be considered a priority risk group to receive the seasonal influenza vaccine as outlined by the Public Health Agency of Canada. The conditions meeting this criterion included chronic respiratory disease, chronic heart disease, chronic kidney disease, chronic liver disease, chronic neurological disease, diabetes (all types), cancer, immunosuppression, dysfunction of the spleen, and/or BMI > 40^41^.

COVID-19 risk perceptions were assessed by 3 statements and each response was recorded using a 6-level Likert scale ranging from Strongly Agree to Strongly Disagree, and Unsure. Perceived effectiveness of public health interventions to control COVID-19 were assessed by 8 questions, the responses to which were recorded using a 5-level Likert scale ranging from Very Effective to Not at all Effective, and Unsure. Respondents’ confidence that they could comply with various public health measures related to COVID-19 were assessed by 7 items and responses were recorded using a 5-level Likert scale ranging from Very Confident to Not at all Confident, and Unsure. Ability to comply with public health measures due to external influences was assessed using a 5 item Likert scale, ranging from Strongly Agree to Strongly Disagree.

Participants were also asked about their use of face masks and their use of public transportation. Adults living with children under the age of 14 years were asked to provide information about childcare provision during school and daycare closures and whether their childcare providers were members of their household. The complete survey is provided in the **Supplementary Materials**.

### Data Analysis

Demographic characteristics of survey respondents were compared with those from the 2016 Canadian Census in order to ensure that the sample population was generally representative of the Canadian population.

Attitudes towards the effectiveness of COVID-19 measures and confidence in individuals’ ability to comply with such measures were aggregated to provide binary measures of agreement (strongly or somewhat); confidence (very or fairly); and perceived effectiveness of measures (very or fairly); with the other category comprised of neutral responses, non-agreement or uncertainty. For a question regarding expectations of coworkers regarding working while ill, the responses “somewhat disagree” and “strongly disagree” were combined to form “Disagree” while all other responses were combined. The proportion of respondents who agreed, were confident, or thought each measure was effective were calculated for each of the questions about attitudes toward COVID-19 public health measures. Chi-square analyses of individual contingency tables were conducted to further explore these data by respondent demographics and household characteristics. The Bonferroni correction was applied for each of the indicators of attitudes toward and ability to comply with public health measures to account for multiple comparisons within each measure. Therefore, a relationship was considered significant if the p-value was less than the corrected value (0.05/19 = 0.0026).

Logistic regression models were developed to identify factors associated with: 1) mask use in the 24 hours prior to survey completion, and 2) reporting direct contact with individuals outside of the respondent’s immediate household in the seven days prior to survey completion. Univariable models were first assessed using a liberal p-value of less than 0.3 to determine eligibility for inclusion in the multivariable models. Variables included in the initial full model for mask use included age, gender, risk group status, size of geographic region of residence, household income, education level, employment status, household composition, household size, as well as two indicators of perceived risk of COVID-19 to self and one indicator of perceived risk of transmission to others. The initial full model assessing factors associated with engaging in an activity with non-household contacts included respondents’ perceived effectiveness of reducing contacts to mitigate transmission in addition to all variables included in the model described above for mask use.

A backward elimination procedure was used to evaluate variables for inclusion in the final multivariable regression models. Confounding was assessed by examining the variables in the model for changes once the potentially confounding variable was excluded from the model. Once the final model was identified, all two-way interaction terms involving age group with the other predictor variables were assessed. Age group was of interest because it was significantly associated with most measures of perceived effectiveness and ability to comply with public health measures. All data were analysed using RStudio Version 1.2.5033 ^42^.

## Data Availability

Data are available upon request from the authors.

## Acknowledgements

G.B and A.L.G. are supported by the Canada Research Chairs program. D.F.N and A.R.T are supported by the Canadian Institutes for Health Research (CIHR). Z.P. is supported by the Natural Sciences and Engineering Research Council (NSERC). E.M. and P.J.L. are supported by Heritage Canada and the Social Sciences and Humanities Research Council (SSHRC). Funding to support data collection was provided by the Public Health Agency of Canada (PHAC), The National Collaborating Centre for Infectious Diseases (NCCID), and the University of Guelph. We thank J. Lau for support in programming the survey. The funders had no role in study design, data collection and analysis, decision to publish or preparation of the manuscript.

